# Immune cells in bronchoalveolar lavage fluid of Ugandan adults who resist versus those who develop latent *Mycobacterium tuberculosis* infection

**DOI:** 10.1101/2021.01.25.21250463

**Authors:** Bonnie A. Thiel, William Worodria, Sophie Nalukwago, Mary Nsereko, Ingvar Sanyu, Lalitha Rejani, Josephine Zawedde, David H Canaday, Catherine M Stein, Keith A Chervenak, LaShaunda L Malone, Ronald Kiyemba, Richard F Silver, John L Johnson, Harriet Mayanja-Kizza, W Henry Boom

## Abstract

**Background:** The search for immune correlates of protection against *Mycobacterium tuberculosis* (MTB) infection in humans is limited by the focus on peripheral blood measures. Bronchoalveolar lavage (BAL) can safely be done and provides insight into cellular function in the lung where infection is first established. In this study, blood and lung samples were assayed to determine if heavily MTB exposed persons who resist development of latent MTB infection (RSTR) *vs* those who develop latent MTB infection (LTBI), differ in the make-up of resident BAL innate and adaptive immune cells.

**Methods:** Bronchoscopy was performed on 21 healthy long-term Ugandan RSTR and 25 LTBI participants. Immune cell distributions in BAL and peripheral blood were compared by differential cell counting and flow cytometry.

**Results:** The bronchoscopy procedure was well tolerated with few adverse reactions. Differential macrophage and lymphocyte frequencies in BAL differed between RSTR and LTBI. When corrected for age, this difference lost statistical significance. BAL CD4+ and CD8+ T cells were almost entirely composed of effector memory T cells in contrast to PBMC, and did not differ between RSTR and LTBI. BAL NKT, γδ T cells and NK cells also did not differ between RTSR and LTBI participants. There was a marginally significant increase (p=0.034) in CD8 T effector memory cells re-expressing CD45RA (T_EMRA_) in PBMC of LTBI *vs* RSTR participants.

**Conclusion:** This observational case-control study comparing unstimulated BAL from RSTR *vs* LTBI, did not find evidence of large differences in the distribution of baseline BAL immune cells. PBMC T_EMRA_ cell percentage was higher in LTBI relative to RSTR suggesting a role in the maintenance of latent MTB infection. Functional immune studies are required to determine if and how RSTR and LTBI BAL immune cells differ in response to MTB.

## Introduction

*Mycobacterium tuberculosis* (MTB) is most frequently transmitted through inhalation of aerosolized bacilli in small droplet nuclei by close contact with a person with pulmonary tuberculosis (TB). Most persons thus infected are thought to develop a latent infection (LTBI) which is reflected in a cellular immune response to MTB antigens measured by tuberculin skin test (TST) or interferon-gamma release assay (IGRA). Most persons with LTBI remain well and only a small proportion progress to TB disease. We and others have found that a small number of heavily MTB exposed persons do not develop a positive TST or IGRA, indicating that they resist developing traditional LTBI. This LTBI resister phenotype remains stable over time and is not attributable to known demographic, clinical or risk factors [1–4]. Immunological comparisons of blood samples from persons with LTBI with those who resist traditional LTBI, i.e. RSTR, indicate that they differ in both adaptive and innate immune responses to MTB [1,5,6]. Since the lungs are the primary portal of entry for MTB in humans, studies of innate and adaptive immune cells in the human lung in LTBI and RSTR are necessary to understand how MTB is controlled in the lungs of LTBIs and RSTRs.

Bronchoalveolar lavage (BAL) has been successfully used for over 30 years to sample the airways of willing participants for clinical research and has been shown to be safe with few complications [7,8]. Studies of pulmonary immune responses in MTB exposed persons are limited but provide some evidence of immune processes that are unique to the lung environment relative to blood and may be enhanced in newly exposed participants [9]. Recently, flow cytometry has been used in conjunction with BAL sampling to study immune cell frequencies associated with TB with the finding that macrophage percentages were decreased in patients with TB relative to healthy participants and those with other lung diseases [10].

In this case-control study we performed bronchoscopy on 21 long-term Ugandan RSTR participants and 25 persons with stable LTBI to test the hypothesis that people who resist traditional LTBI have a local enrichment of innate and/or T-cell subsets in the lung that contribute to control of MTB. BAL and peripheral blood mononuclear cell (PBMC) numbers and percentages were compared between LTBIs and RSTRs to look for immunologic markers associated with the resistant phenotype.

## Materials and Methods

### Study participants

Bronchoscopy participants were healthy, nonsmoking, 18–49 year-old, HIV-uninfected Ugandan adults who previously participated in a long-term follow-up study of household contacts of a person with pulmonary TB [4]. In brief, RSTR and LTBI participants were followed for a mean of 13 years and classified as having the resistance phenotype defined by at least 2 negative tuberculin skin tests (TST) 48 to 72 hours after intradermal injection of 5 TU of PPD-S, (Sanofi Pasteur TUBERSOL, Toronto Ontario Canada) and at least 2 negative IGRA tests (Quantiferon TB Gold Assay (QFT), Quiagen/Cellestis, Germantown, MD, USA) performed during the 2 year long-term follow-up period. The QFT was interpreted according to the manufacturer’s instructions. Participants were classified as stable LTBI if all follow-up TST and IGRA tests were positive during the 2 year long-term follow-up study and they remained asymptomatic for TB. The enrollment period was between December 2017 and January 2020. All re-contacted participants for this study had their MTB infection status reconfirmed by QFT at the time of bronchoscopy, and were required to have a normal chest radiograph done within the previous 6 months, a normal (or out-of-range and not clinically significant) complete blood count, electrolyte, renal and hepatic function tests, and normal prothrombin and activated partial thromboplastin time for enrollment.

Persons with a history of asthma or other chronic lung disease, cardiac disease, cancer, seizure disorder, hypertension, diabetes mellitus, a history of adverse reactions to lidocaine, or prior TB were excluded. Participants with any signs or symptoms of active TB, a chest radiograph suspicious for active TB, or who were pregnant or lactating were also excluded. The participant sample is representative of the larger population of healthy Ugandan TB contacts. The study protocol was reviewed and approved by the institutional review boards of the Makerere University School of Biomedical Sciences and University Hospitals Cleveland Medical Center. All participants gave written informed consent for study participation. Participant demographic information is available in Table 1 in the S1 Dataset.

**Table 1.**
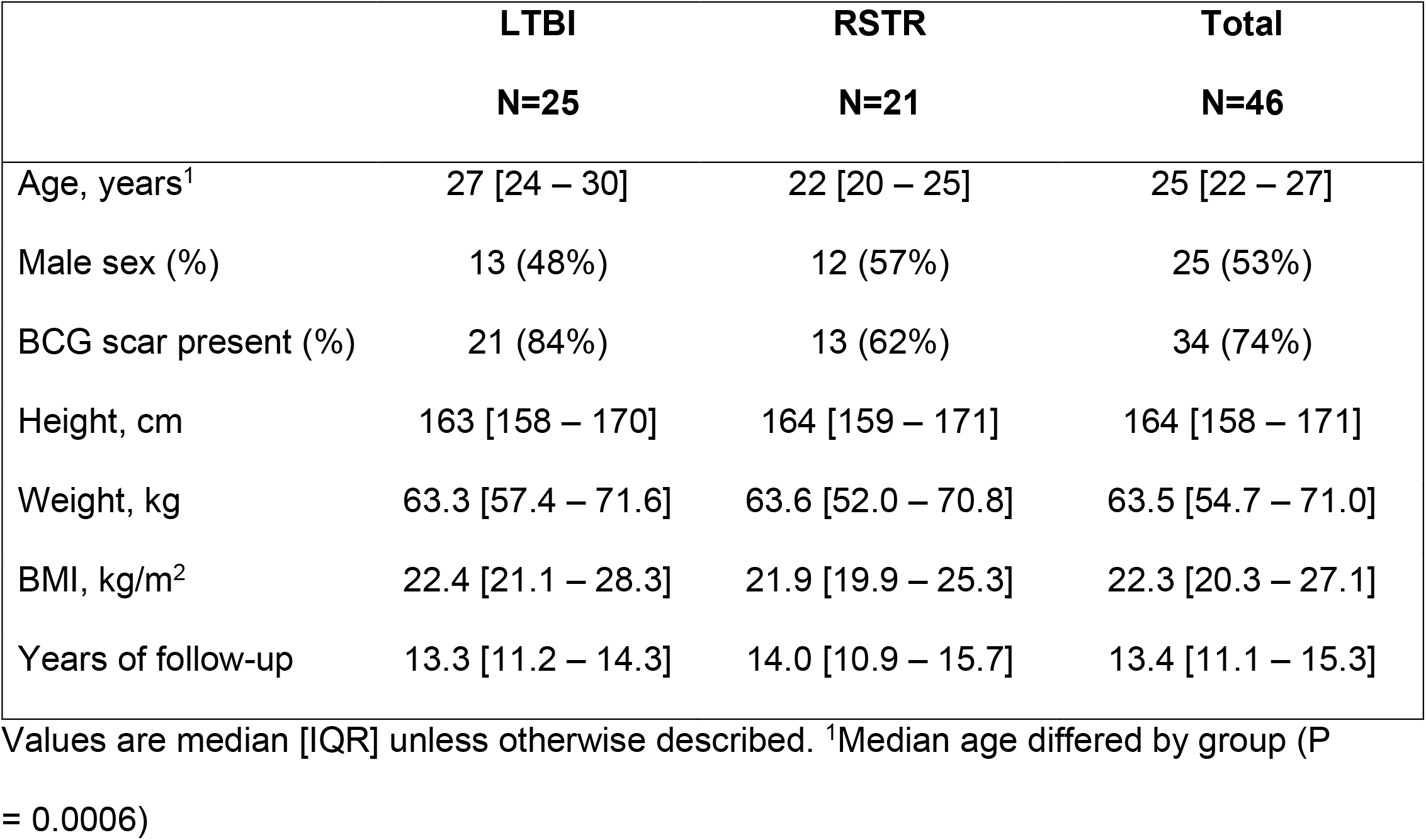
Characteristics of study participants

|  | **LTBI** | **RSTR** | **Total** |
| --- | --- | --- | --- |
|  | **N=25** | **N=21** | **N=46** |
| Age, years<sup>1</sup> | 27 [24 – 30] | 22 [20 – 25] | 25 [22 – 27] |
| Male sex (%) | 13 (48%) | 12 (57%) | 25 (53%) |
| BCG scar present (%) | 21 (84%) | 13 (62%) | 34 (74%) |
| Height, cm | 163 [158 – 170] | 164 [159 – 171] | 164 [158 – 171] |
| Weight, kg | 63.3 [57.4 – 71.6] | 63.6 [52.0 – 70.8] | 63.5 [54.7 – 71.0] |
| BMI, kg/m<sup>2</sup> | 22.4 [21.1 – 28.3] | 21.9 [19.9 – 25.3] | 22.3 [20.3 – 27.1] |
| Years of follow-up | 13.3 [11.2 – 14.3] | 14.0 [10.9 – 15.7] | 13.4 [11.1 – 15.3] |

Values are median [IQR] unless otherwise described. <sup>1</sup>Median age differed by group ( $P = 0.0006$ )

### Bronchoscopy and bronchoalveolar lavage

All bronchoscopies were performed under topical anesthesia via a trans-nasal approach in the bronchoscopy unit at Naguru Hospital, Kampala, Uganda. Pre-procedure lung and cardiac examinations were normal for all participants. Participants received nothing by mouth for at least 6 hours before bronchoscopy. Before each procedure, participants received aerosolized 2% lidocaine and gargled several times with 2% lidocaine solution for 30 seconds. Topical anesthesia of the nasal passage was performed using 2% viscous lidocaine jelly applied with cotton applicators. Further local anesthesia was provided by topical application of 1 to 2 ml aliquots of 1% or 2% lidocaine to the airways via the bronchoscope. The total dose of lidocaine administered for local and airway anesthesia was 7 mg/kg (maximum 400 mg). Participants received supplemental oxygen (2 l/min) via nasal cannula and all vital signs were monitored using a bedside monitor during bronchoscopy. Bronchoalveolar lavage was performed by wedging the tip of the bronchoscope in a subsegment of the right middle lobe or lingua. Aliquots of 30 ml of ambient temperature sterile normal saline solution for IV infusion (up to a total of 240 ml) were instilled into the airway and gently aspirated using a 60 ml syringe. All participants were observed in the bronchoscopy unit for at least 120 min after the procedure. A blood sample from all participants was obtained for parallel assessments of immune cells on the day of bronchoscopy. Subjects were provided with contact information for the study coordinator and one of the investigators, and were advised to call if any symptoms of concern arose. All participants were seen at a post-procedure visit within 72 hours of research bronchoscopy, and then followed up further, if needed, until resolution of any post-bronchoscopy symptoms.

### Processing of bronchoalveolar lavage fluid and whole blood

The syringes containing the aspirated BAL fluid were labelled, placed on ice blocks, and transported promptly to the laboratory using a preconditioned OrcaTherm cooler (Intelsius, York, UK). An aliquot was taken from the first syringe and sent to the microbiology laboratory for MTB culture. All BAL samples were determined to be MTB culture negative. Fluid was then aliquoted into 50 cc polypropylene tubes, and the total volume of BAL fluid recovered recorded.

Peripheral blood mononuclear cells were isolated from whole blood using Ficoll-Paque PLUS (GE Healthcare) density gradient and Leucosep separation tubes (Greiner Bio-One, Kremsmunster, Austria). The gradient is centrifuged for 10 min at 1000 x g at room temperature. The resulting PBMC layer was decanted, washed 2x and counted. Individual BAL cell yields and BAL cell differentials are available in Table 2a and Table 2b in S1 Dataset.

**Table 2.** BAL fluid recovery, cell yield and cell differentials of LTBI and RSTR participants

|  | LTBI | RSTR | P-value | Age adjusted P-value |
| --- | --- | --- | --- | --- |
| BAL fluid | N=25 | N=21 |  |  |
| Lavage fluid recovered, ml | 177 [169 - 193] | 186 [177 - 191] | 0.33 | 0.40 |
| (% total volume) | (74%) | (78%) |  |  |
| BAL cell number ($\times 10^6$)* | 14.2 [12.8 - 21.0] | 14.6 [9.6 – 21.0] | 0.77 | 0.50 |
| BAL cell number (x10<sup>4</sup>)/ml fluid recovered* | 8.0 [6.8 – 11.1] | 7.7 [5.5 – 11.0] | 0.49 | 0.26 |
| Viability (% live cells)* | 91.5 [85.1 – 93.9] | 91.1 [85.5 – 95.0] | 0.98 | 0.31 |
| BAL cell differential | N = 23 | N = 20 |  |  |
| % Macrophages | 80.5 [76.2 – 86.7] | 87.8 [84.0 – 91.1] | 0.02 | 0.25 |
| % Lymphocytes | 13.5 [10.7 – 15.7] | 10.5 [7.3 – 12.4] | 0.05 | 0.70 |
| % Neutrophils | 2.0 [1.0 – 3.2] | 1.5 [0.4 – 2.5] | 0.09 | 0.10 |
| % Eosinophils | 1.0 [0.5 – 2.2] | 0.5 [0.0 – 1.5] | 0.06 | 0.47 |
| % Basophils | 0.5 [0.0 – 1.5] | 0.0 [0.0 – 0.0] | 0.007 | 0.18 |

Values are median [IQR]. \*Nine participants in the LTBI group and 4 participants in the RSTR group were missing values for total cell number and viability. Two LTBI participants and 1 RSTR participant had missing differential counts for technical reasons. P-values are from a Mann-Whitney-Wilcoxon test of median differences. Age adjusted P-values are from a fitted linear model. NS=not significant.

### Measurement of BAL and PBMC viability and cell type

BAL samples were immediately centrifuged at 300 x g for 10 min at room temperature (18-22^0^C). The supernatant was harvested, aliquoted and stored. BAL cell pellets were gently loosened and cells combined into one 50ml falcon tube. BAL cells were resuspended in 10ml of cold medium (10% Fetal Bovine serum + 1% L-Glutamine + 1% Penicillin in RPMI). Cells were stained with trypan blue for counting and assessment of viability.

Cytospin slides were prepared using 30,000 and 60,000 cells from each BAL sample in duplicate. Resuspended BAL cells were loaded onto the slides by placing the cell volume into a slide centrifugation apparatus (Cytofunnel, #5991040; Shandon, Pittsburgh, PA) followed by centrifuging at 800 x g for 8 min. Slides were air dried and stained with a rapid Wright-Giemsa stain method (LeukoStat, #C430; Fisher Diagnostics, Pittsburgh, PA). Cell differentials were determined by counting 100 cells from each sample under 100X objective by light microscopy.

### Flow cytometric analysis of BAL and PBMC T cell populations

BAL cells (200,000 – 500,000 cells) and PBMC (100,000 – 200,000) were resuspended in Facs Buffer (10%FBS+4mM EDTA +PBS) and stained for 20 minutes at room temperature with the following antibodies used per manufacturer’s instructions: CD3-FITC (Biolegend, #344804), CD4-APC-Cy7 (BD Biosciences, #BD557871), CD8-APC (Biolegend, #344722), CD56-BV421 (Biolegend, #362552), Gamma-delta-PE (Biolegend, #331210), CCR7-PECY7 (Biolegend, #353226) and CD45RA-PECY5 (Biolegend, #304110). Compensation was performed using antibody capture beads (anti-mouse k; BD Biosciences, #552843). Zombie Aqua fixable viability kit (BioLegend, #433101) was used as the live/dead discriminator. Cells were fixed, acquired and analyzed on a MacsQuant flow cytometer using MacsQuant software version 2.11.176.19438. Analysis was performed using Flow Jo version 10.6 (Oregon, USA). Cells where gated on lymphocyte sized, singlet, live cells. BAL and PBMC flow cytometric counts and percentages for each participant can be found in the S2 Dataset.

### Statistical analysis

Means were compared between donor groups using Welch’s two-sample t-test. Medians were compared using the Mann-Whitney-Wilcoxon test. Fisher’s exact test was used for testing independence of count data. A general linear model was used to evaluate cell percentage as a function of phenotype class with age as a covariate.

Correlation coefficients were estimated by the Spearman method and are notated as r_s_. All statistical analyses were done in R version 3.6.2 (R Core Team, 2020). The data collection and management for this paper was performed using the OpenClinica open source software, version 3.13 (Waltham, MA)

## Results

### Enrollment and clinical characteristics of RSTR and LTBI participants

A pool of 147 HIV negative adults enrolled in a previous longitudinal study and classified as MTB latently infected (LTBI) or resistant to MTB infection (RSTR) were invited to participate in this bronchoscopy study [4]. Seventy-three eligible participants consented, 64 were enrolled and 50 participants (24 RSTR and 26 LTBI) underwent the bronchoscopy procedure. Reasons for screening/enrollment failures included laboratory values out of range (n=11), positive HIV test (n=1), positive pregnancy test (n=1), chronic or comorbid illness (n=2), age out of range (n=1), and failure to arrive for the pre-bronchoscopy or bronchoscopy visit (n=12). There were no serious adverse reactions to the bronchoscopy procedure. Sore throat (n=7), chest pain (n=1) and fever(n=1) were the only related adverse events. Non-related events included lower back pain (n=1), flu (n=1), elevated blood pressure (n=1) and jaw pain (n=1).

At the time of bronchoscopy, all participants had their infection status re-tested by IGRA. Twenty-one of the 24 resister participants remained QFTG negative and 3 participants converted to IGRA positive. One LTBI participant had an uninterpretable IGRA result. The final dataset included 21 RSTR and 25 LTBI participants.

The median age was 25 years old and 53% of participants were males. Table 1 shows a comparison of demographic and clinical parameters and total long-term follow-up period for LTBI and RSTR participants. The median age in the RSTR group (22 years) was less than median age of LTBI participants (27 years) (P = 0.0006); otherwise, there were no differences between groups.

### BAL fluid recovery, cell yield and cell differentials of LTBI and RSTR participants

Although participants were all non-smokers, BAL pellets were darkly stained indicating exposure to carbon pollutant particles in Urban Kampala. There were no differences in pellet staining between RSTR and LTBI groups. Table 2 summarizes the BAL fluid yield and cell type comparisons between groups. LTBI and RSTR participants had similar BAL yields in terms of BAL volume and cell recovery per ml of fluid. The median percentage of macrophages was higher in RSTR (87.8%) *vs* LTBI participants (80.5%) but this difference lost significance after taking age into account. Age was inversely correlated with macrophage percentage (r_s_ = −0.47, P=0.001) and was higher in LTBI participants, therefore inclusion of age in the linear model reduced the effect of infection status. Lymphocyte percentage was positively correlated with age (r_s_ = 0.46, P=0.001). Neutrophil, eosinophil and basophil percentages were not significantly different between participant groups after adjusting for age.

### NK and T cell subsets in BAL cells and PBMC from RSTR and LTBI participants

A subset of 27 participant BAL cell and PBMC samples were analyzed by flow cytometry for T subsets and NK cells. Lymphocyte single cells were sub-setted into CD3 positive and negative cells. CD3 negative cells were gated for CD56+NK cells while CD3+ cells were gated into CD4+, CD8+, γδ and CD56 positive NK T cells. Fig 1 shows the gating strategy.

**Fig 1:**
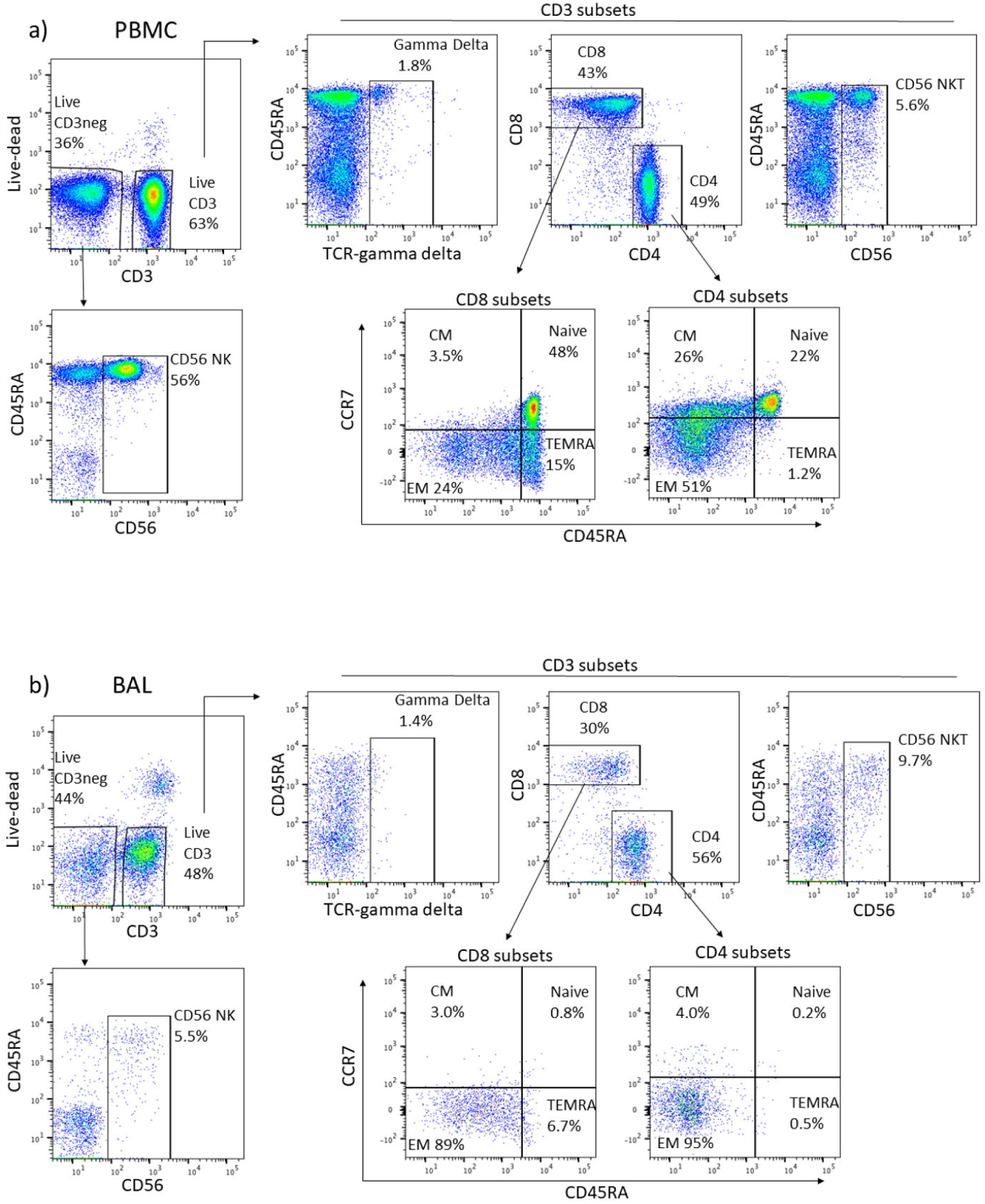
A representative flow analysis for a) PBMC and b) BAL cells. CD56 NK percentage was quantified as a subset of the live CD3-population. Live CD3 lymphocytes were gated as shown to determine gamma delta, CD56 NKT, CD8, and CD4 subsets. CD45RA and CCR7 markers were then used to subset CD4 and CD8 cells into CM, EM, Naive and TEMRA groups.

Major cell subset percentages in PBMC and BAL within LTBI and RSTR cohort are shown in Table 3. There were no significant differences between RSTR and LTBI group medians in BAL fluid recovery, cell yield and cell differentials of LTBI and RSTR participants in this initial analysis.

**Table 3:** CD3+ T cell subset and CD3- NK cell percentages in BAL and PBMC lymphocytes from LTBI and RSTR participants

|  | BAL |  | PBMC |  |
| --- | --- | --- | --- | --- |
| Phenotype (flow) | LTBI<br>N=11*<br>% [range] | RSTR<br>N=14<br>% [range] | LTBI<br>N=13<br>% [range] | RSTR<br>N=14<br>% [range] |
| CD4+ T cell | 55.5<br>[45.2 – 61.6] | 54.1<br>[42.5 – 63.8] | 54.2<br>[49.8 – 57.0] | 58.8<br>[50.3 – 62.1] |
| CD8+ T cell | 32.4<br>[25.6 – 37.8] | 32.6<br>[26.8 – 50.2] | 33.1<br>[30.7 – 38.4] | 30.0<br>[23.2 – 31.9] |
| CD56+ NK T cell | 4.1<br>[3.3 – 6.3] | 6.0<br>[3.2 – 8.7] | 6.6<br>[4.4 – 13.1] | 4.9<br>[2.5 – 8.4] |
| $\gamma\delta$ T cell | 1.2<br>[0.6 – 2.2] | 2.3<br>[1.6 – 3.2] | 1.5<br>[0.9 – 3.4] | 2.2<br>[1.3 – 4.0] |
| CD56+ NK cell | 4.9<br>[2.9 – 7.2] | 4.4<br>[1.8 – 12.3] | 27.7<br>[14.7 – 38.3] | 12.6<br>[6.5 – 40.3] |

NK=Natural Killer. CD4+, CD8+, CD56+ and $\gamma\delta$ were calculated as a percentage of CD3+ lymphocytes, CD56+ NK cells were calculated as a percentage of the parent CD3- lymphocyte population. \*2 LTBI participants were missing BAL flow results for technical reasons.

### CD4+ and CD8+ T cell subsets in BAL and PBMC from RSTR and LTBI participants

Next, the distribution of CD4+ and CD8+ T cell subsets were determined in BAL cells and PBMC. Previous studies have found differences in unstimulated PBMC T cell subset percentages when persons with LTBI were compared to healthy MTB-uninfected persons [10,11]. Using CD45RA and CCR7 as markers, naïve T cells (T_N_ = CCR7+/CD45RA+), effector memory (T_EM_ = CCR7-/CD45RA-), central memory (T_CM_ = CCR7+/CD45RA-), and effector memory re-expressing CD45RA (T_EMRA_ = CCR7-/CD45RA+) were identified by flow cytometry.

BAL CD4+ and CD8+ T cell subsets were differentiated by CCR7 vs CD45RA as shown in Fig 2a. Median percentages did not differ significantly between RSTR and LTBI participants after adjustment for age. The same subsets were analyzed in PBMC obtained at time of BAL as shown in Fig 2b. Interestingly, CD8+ CCR7-/CD45RA+ T cells (T_EMRA_) were significantly higher in LTBI compared to RSTR participants and remained significant after adjustment for age (P = 0.034). CD8+ T_EMRA_ cell % did not correlate with peripheral blood lymphocyte count (data not shown).

**Fig 2:**
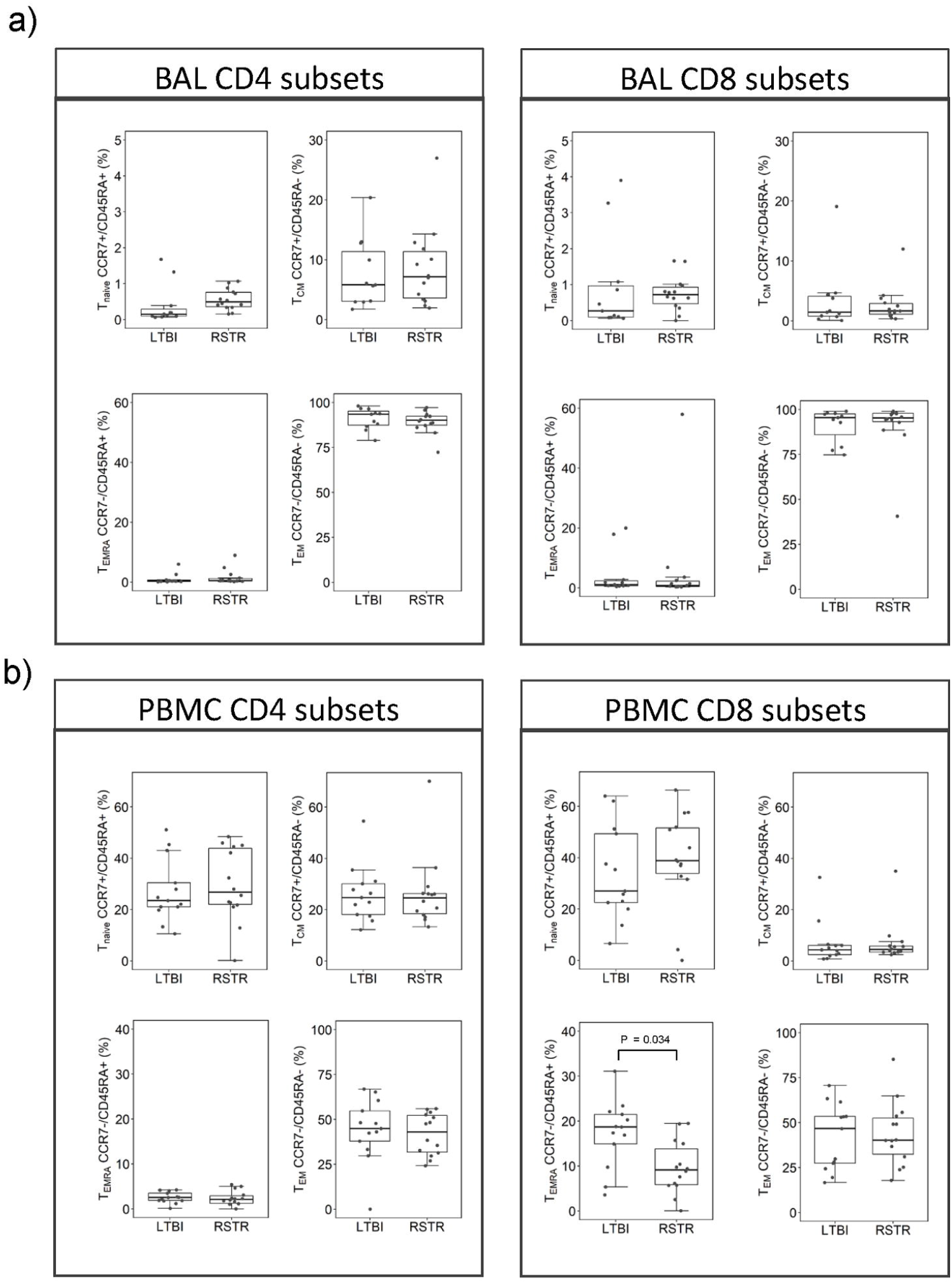
CD4+ and CD8+ cell subset percentages based on CD45RA and CCR7 gating. a) BAL lymphocyte subsets: N=11 LTBI and14 RSTR, b) PBMC lymphocyte subsets: N=13 LTBI and14 RSTR. PBMC CD8 T_EMRA_ median percentage was larger in LTBI compared to RSTR (unadjusted P=0.02/adjusted P=0.034). Comparison of BAL vs PBMC: T_EM_ median CD4+ and CD8+ percentages were larger in BAL compared to PBMC (P < 0.0001).

When T cell subsets were compared between BAL and PBMC from the same participant, BAL CD4+ and CD8+ T cells had higher median frequencies of T_EM_ cells compared to PBMC CD4+ and CD8+ respectively (P < 0.0001). Consistent with this finding was a decrease in BAL of naïve CD4+ and CD8+ (P < 0.0001), T_EMRA_ (CD4+ P=0.016, CD8+ P < 0.0001) and T_CM_ (CD4+ P <0.0001, CD8+ P=0.004) compared to PBMC.

## Discussion

In previous studies we and others have demonstrated that there is a small but significant number of heavily MTB exposed persons who remain healthy and do not develop a positive TST and/or IGRA response which is diagnostic for LTBI. We have labeled these persons LTBI resisters, i.e. RSTR. We have found in our studies in Uganda that these RSTRs have unique peripheral blood T cell and antibody responses to MTB, indicating sensitizing and likely controlled MTB infection using non-traditional LTBI-like responses [6]. Since TB and MTB infection affect primarily the lung, we set out to determine the feasibility and safety of research bronchoscopy in Uganda and analyze the baseline BAL immune profile of long-term stable RSTRs and LTBIs (Table 1).

We found that research BAL using local anesthesia was safe and well-tolerated in our 21 Ugandan RSTRs and 25 LTBIs. Analysis of baseline BAL and PBMC immune cell distribution by cytology and flow cytometry found subtle differences. RSTR had decreased percentage of lymphocytes and increased macrophages compared to LTBIs, as well as very minor difference in basophils. In light of Young, et al [10] suggesting that age may impact BAL cell distribution, we corrected for the small age difference between RSTR and LTBI (22 and 27 years, respectively). This negated the statistical difference we initially observed. Future studies with a larger sample size and age range will determine if there is a difference between RSTRs and LTBIs in baseline macrophage numbers.

In response to antigen, naïve CD4+ and CD8+ T cells differentiate into long-lived memory subsets. Effector memory T_EM_ and T_CM_ subsets circulate in blood, ready to respond to antigen at re-exposure. In BAL we found very few naïve and a predominance of effector/memory CD4+ and CD8+ T cells. While this has been described for CD4+ T cells, few studies have commented on the predominance of effector/memory CD8+ T cells. Due to prolonged and stable MTB infection in LTBI, we hypothesized that this group might have a greater number of T memory cells relative to RSTR. Our results show no differences in CD4+ and CD8+ T memory cell percentages between LTBI and RSTR participants in either PBMC or BAL. However, we did find a slightly higher percentage of PBMC CD8+ T_EMRA_ cells in LTBI participants relative to RSTRs (Fig 2b). This difference remained significant after adjusting for age. Interestingly, T_EMRA_ cells can be the target cells of anti-TNFα therapy, resulting in an increased risk of MTB reactivation in LTBI [13]. This result requires further validation but suggests that long-term stable LTBI is associated with an increase in CD8+ T_EMRA_ cells.

A limitation of our study was its power to detect significant differences between cell subsets due to the large variances between participants, which could be overcome by a larger sample size. In addition, we did not compare MTB antigen-specific cell frequency differences in RSTR and LTBI participants. Studies comparing MTB specific antigen responses in patients with active TB vs controls have found significant differences among CCR7/CD45RA lymphocyte subsets in blood [14,15]. MTB specific antigen responses in BAL from participants in a non-endemic setting showed that TST-positive donors harbored IFN-gamma producing CD4+ T-cells in response to PPD stimulation at baseline while TST-negative donors did not [16,17]. Antigen-specific responses in CD4+ and other T cell subsets in the lung may identify clear differences in RSTR vs LTBI lung T cell responses to MTB.

Although LTBI is the most common outcome after prolonged exposure to MTB, some individuals clear the infection through innate immunity or control MTB through alternative adaptive immune mechanisms in the lung. This study provides an initial look into BAL and PBMC cell frequencies in RSTR and LTBI groups, and adds to experience about the safety and feasibility of research bronchoscopy in resource limited and TB endemic settings such as Uganda. In the unstimulated baseline condition, we found no major differences in immune cell distributions in BAL and PBMC of RSTR and LTBI. We found preliminary evidence of increased percentage of CD8 T_EMRA_ cells in the PBMC compartment. Future studies will include phenotyping of BAL and PBMC immune cells in response to MTB bacilli and antigens to determine what differences in immune responses to MTB exist in the lungs of MTB exposed persons with latent infection compared to resisters.

## Data Availability

The data referred to in the manuscript are available in the supplemental files.

## Acknowledgements

We acknowledge the contributions made by senior physicians, medical officers, nurses, health visitors, laboratory and data personnel: Drs Alphonse Okwera, Moses Joloba and Brenda Okware, Dorcas Lamunu, Deborah Nsamba, Annet Kawuma, Saidah Menya, Joan Nassuna, Joy Baseke, Hussein Kisingo, Michael Odie, Racheal Nakyesige, Joanita Nassali, Anna-Ritah Namuganga and Jessica Walrath. This study would not be possible without the generous participation of the Ugandan patients and families.

## Supporting Information

**S1 Dataset. Participant demographics, BAL cell yields and BAL cell differentials**.

**S2 Dataset. BAL and PBMC flow cytometry cell counts and percentages**.

